# Uncovering COVID-19 Transmission Tree: Identifying Traced and Untraced Infections in an Infection Network

**DOI:** 10.1101/2024.05.01.24306730

**Authors:** Hyunwoo Lee, Hayoung Choi, Hyojung Lee, Sunmi Lee, Changhoon Kim

## Abstract

We present a comprehensive analysis of COVID-19 transmission dynamics using an infection network derived from epidemiological data in South Korea, covering the period from January 3, 2020, to July 11, 2021. This network, illustrating infector-infectee relationships, provides invaluable insights for managing and mitigating the spread of the disease. However, significant missing data hinder the conventional analysis of such networks from epidemiological surveillance. To address this challenge, our research suggests a novel approach for categorizing individuals into four distinct groups, based on the classification of their infector or infectee status as either traced or untraced cases among all confirmed cases. Furthermore, the study analyzes the changes in the infection networks among untraced and traced cases across five distinct periods. The four types of cases emphasize the impact of various factors, such as the implementation of public health strategies and the emergence of novel COVID-19 variants, which contribute to the propagation of COVID-19 transmission. One of the key findings of this study is the identification of notable transmission patterns in specific age groups, particularly in those aged 20–29, 40–69, and 0–9, based on the four type classifications. Moreover, we develop a novel real-time indicator to assess the potential for infectious disease transmission more effectively. By analyzing the lengths of connected components, this indicator facilitates improved predictions and enables policymakers to proactively respond, thereby helping to mitigate the effects of the pandemic on global communities.

## 1 INTRODUCTION

COVID-19, caused by the severe acute respiratory syndrome coronavirus 2 (SARS-CoV-2), was declared a pandemic by the World Health Organization on March 11, 2020. According to the World Health Organization’s weekly epidemiological update released on February 2, 2021, the epidemic of COVID-19 spread rapidly to more than 200 countries. Without effective control measures, the rapidly increasing number of COVID-19 cases will greatly increase the burden of clinical treatments. This situation may lead to a critical shortage of healthcare system capacity for severe cases, ultimately resulting in a sharp and alarming increase in mortality rates. Consequently, various control measures were implemented, leading to observed fluctuations in the efficacy of strategies like contact tracing and isolation of confirmed cases throughout the pandemic (1). South Korea, first reporting its COVID-19 case on January 19, 2020 (2, 3), has experienced multiple waves of outbreaks, in response to which it actively implemented control measures such as social distancing, mask-wearing, lockdowns, and enhanced efforts in testing and contact tracing. Especially, active contact tracing has generated significant epidemiological data, enabling analysis of extensive infection networks (4). Understanding the infection network for COVID-19 is crucial for several reasons. First and foremost, it allows us to grasp the dynamics of the virus’s transmission within a population (5). By mapping out how individuals infect each other, we gain valuable insights into the patterns and pathways through which the virus spreads (1). Additionally, studying the infection network aids in the identification of key factors influencing the transmission (2). This includes factors such as age-specific patterns, which can help tailor public health measures to specific demographics, ultimately improving the effectiveness of containment strategies (6).

Previous research focused on cluster analysis, reproduction number, and network analysis to address key transmission factors and assess the effectiveness of various interventions during COVID-19 pandemic (3, 6, 7, 8, 9, 10, 11, 12). In (7, 8) authors investigated COVID-19 transmission by age group, aiding in identifying the primary age groups fueling the spread and formulating age-specific response strategies. It scrutinized the infection spread by clusters, offering insights into evaluating social distancing measures outlined in (3, 6, 9). Examining cluster type frequency in both the initial and subsequent epidemic waves enables the development of an effective strategy for controlling outbreaks (3). Network analysis facilitates assessing specific vertices’ importance and understanding the relationships between them (2, 5, 13). Furthermore, Wang et al.(10) and Zhang et al.(11) investigated the basic reproduction number R_0_ of COVID-19, which represents the transmission potential of an infectious disease in the early phase of an epidemic (12). The time-dependent reproduction number R_*t*_ represents the instantaneous reproduction number, indicating the expected number of secondary infections caused by an infector at a specific point in time (12).

In the context of COVID-19 policies, our current knowledge of how infections spread through transmission networks is primarily based on virtual data and theoretical models (14, 15), with evidence from actual data (16, 17, 18) being limitedly available. The infection network generated from actual epidemiological data contains numerous missing data, resulting in many connected components, creating a disparity from analyses based on virtual data. Contact tracing is commonly recommended for controlling COVID-19 outbreaks, yet its effectiveness is unclear. Studies evaluating the effectiveness of contact tracing are categorized into observational studies (19, 20, 21, 22) and modeling studies (1, 23, 24, 25). Our study suggests that analyzing the classification of four types of confirmed cases in the infection network, along with the distribution of connected component lengths, can broaden insights into contact tracing and dynamics of disease transmission. A pivotal study analyzing changes in the infection pattern structure between infectors and infectees based on age groups (26) is also essential. Surprisingly, there has been no previous study on this specific topic for COVID-19 infection between infectors and infectees in South Korea.

This paper is motivated by the recognition of differences in infection networks generated from actual data versus virtual data. This research has established an infection network by assigning an infector to all infectees from actual epidemiological data KDCA (27) from January 3, 2020, to July 11, 2021, in South Korea. It is shown that the established infection network comprises many connected components due to missing vertices (individuals) and edges (infection events). Consequently, we proposed a method of categorizing individuals as either (i) infectors, who are aware of the infectees they have transmitted the virus to, or (ii) infectees, who are cognizant of their infector. This method allows for the categorization of vertices in the numerous distinct connected components from a common perspective and facilitates the derivation of analysis for each vertex. Furthermore, several properties were established from the method. This paper analyzed the infection network in terms of time and age groups using a four-type categorization method and proposes a new real-time calculated indicator of infectious disease transmission potential. Next, the indicator was compared with the Cori reproduction number R_*t*_ (12). Age groups are evenly distributed into nine categories, up to 90 years old. To characterize each wave, the period is divided into five phases, accounting for epidemic control measures and the progression of epidemic waves.

Our analysis focuses on the comprehensive infection network across age groups, revealing how infection spread patterns evolve over time, and concentrates on methods to obtain meaningful information in the presence of substantial missing data. This analytical approach, based on epidemiological data, emphasizes the role of active contact tracing by governments. Ultimately, our research suggests that active contact tracing in real pandemic situations can offer policymakers data-driven insights for establishing more effective responses, thereby mitigating the pandemic’s impact on global communities.

## 2 METHODS

### 2.1 Data and measurement

#### A Data description

We utilize the COVID-19 data (27) provided by the Korea Disease Control and Prevention Agency (KDCA) from January 19, 2020, to July 11, 2021, to construct the infection network for COVID-19 transmission. In this paper, we analyze the dataset containing 169,597 confirmed cases (real-time reverse transcription polymerase chain reaction test positive cases), focusing on four specific records as follows.

**(ID, age, date of report, ID of the infector)**

Here, the “ID” stands for the identity of the traced infectee, and “age” refers to the infectee’s age. If “ID of the infector” is not traced (untraced), it is assigned a value of 0. Each confirmed case is assigned an anonymized ID number ranging from 1 to 169,146 associated with age, which ranges from 0 to 128, the date of report, and the ID number of the infector. Remark that in general the date of the report may not be exactly the same as the date of infection. The date of the from January 19, 2020, to July 11, 2021.

#### B. Defining five periods of COVID-19 progression

The entire period was segmented into five distinct periods to observe the evolution of infection characteristics. This segmentation considered several critical factors like the emergence of new variants, vaccine rollout, change of social distancing levels, and other intervention measures (28).

- *P* 1 (January 19, 2020 ∼ April 29, 2020): Since the first confirmed COVID-19 case on January 19, 2020, South Korea experienced a moderate rise in cases, peaking at about 694 on February 26, 2020, primarily in Daegu-Gyeongbuk due to a church-related outbreak. Despite subsequent outbreaks at another church and a Seoul call center, daily cases gradually declined. Measures like the first social distancing period (March 22 to April 7, 2020) and a ban on gatherings in entertainment venues (April 8 to April 19, 2020) were enacted, resulting in an average of 145 daily confirmed cases during these periods.
- *P* 2 (April 30, 2020 ∼July 14, 2020): During this period, there was the lowest number of daily confirmed cases compared to other periods. The average number of daily confirmed cases was 37.
- *P* 3 (July 15, 2020 ∼ October 12, 2020): The second epidemic wave in South Korea started with a major outbreak at a Seoul church, accounting for 12% of the total infections in period *P* 3, and was further exacerbated by a large rally on August 15 contributing to 6% of infections. In response, the government escalated Seoul’s social distancing to level 2 on August 16, expanded it nationwide on August 23, and then increased it to level 2.5 in the metropolitan area by August 30. The peak of this wave was on August 24, 2020, with 418 cases, and the average daily confirmed cases during this period was 125.
- *P* 4 (October 13, 2020 ∼February 25, 2021): On October 12, the social distancing level was eased to level 1. *P* 4 coincides with the third epidemic wave, and it started with a gradual increase in daily confirmed cases without any apparent major events. The third epidemic wave peak occurred on December 23, 2020, with 1206 cases. The government raised the social distancing level on December 1 and then again on December 8 and increased screening clinics. During this period, the average number of daily confirmed cases was 463.
- *P* 5 (February 26, 2021 ∼ July 11, 2021): South Korea began its vaccination campaign on February 26, 2021, and then saw an increase in delta variant cases starting April 18, 2021. During this period, the average number of daily confirmed cases was 571.

### 2.2 Infection network of infector and infectee

Network, also called graph mainly in mathematics, has been used as an explanatory tool to describe the dynamics of disease transmission (29). The terms “individuals (confirmed cases)” and “contacts (infects)” in epidemiology can be considered as “vertices” and “edges” in graph theory, respectively. For more details on network epidemiology, see the review (30, 31) and references therein.

Denote the set of all confirmed IDs from January 19, 2020 to July 11, 2021 as, and let the set of all infection events (*m*_−1_, *m*_0_) for the infector *m*_−1_∈ I and its infectee *m*_0_ ∈ I as E. We consider the directed network *G* = (*ℐ,ε*) as an infection network. For complete sampling, the infection network *G* must be weakly connected (replacing all its directed edges with undirected edges produces a connected undirected graph). However, due to the existence of unreported infection cases, it is natural to assume that the network is constructed by incomplete sampling of all confirmed individuals in a population (missing vertices) and incomplete sampling of infection events between individuals (missing edges). So the infection network *G* generated by real data consists of many weakly connected (or just connected components in this paper) due to many missing vertices and edges, i.e., unreported individuals and infections. Hence analysis of unreported infections is crucial for a better understanding of the real infection network in South Korea and other countries.

### 2.3 Four type classifications

Each polymerase chain reaction (PCR)-confirmed case *m*_0_ can be classified into four different types based on (i) as an infector *m* _1_, whether the infectees they have transmitted the virus to have been traced or (ii) as an infectee *m*_1_, whether they are aware of their infector being traced.

i. An individual *m*_0_ ∈ I is said to be “**untraced**-**untraced**” type, denoted by **u-u**, if {*m*_0_ ∈ I|(*m*_−1_, *m*_0_)∈ *ε*}=∅ and { *m*_0_∈ *ℐ*| (*m*_0_, *m*_1_) ∈ *ε*}= ∅, i.e., its infector is missing (untraced) and its infectee is missing or does not exist. Such an individual is represented as an isolated vertex on the network.
ii. An individual *m*_0_ is said to be “**traced-untraced**” type, denoted by **t-u**, if {*m*_0_ ∈ I|(*m*_−1_, *m*_0_)∈ *ε*}=∅ and { *m*_0_∈ *ℐ*| (*m*_0_, *m*_1_) ∈ *ε*}= ∅, i.e., its infector is confirmed (traced) but its infectee is missing or does not exist. Such an individual is represented as a leaf of a directed tree graph.
iii. An individual *m*_0_ is said to be “**untraced-traced**” type, denoted by **u-t**, if {*m*_0_ ∈ *ℐ*|(*m*_−1_, *m*_0_) ∈ E} = ∅ and {*m*_0_ ∈ *ℐ*|(*m*_0_, *m*_1_) ∈ *ε*} ∅, i.e., its infector is not confirmed but its infectee is confirmed. Such an individual is represented as a root of a directed tree graph.
iv. An individual *m*_0_ is said to be “**traced-traced**” type, denoted by **t-t**, if {*m*_0_ ∈ *ℐ*|(*m*_−1_, *m*_0_) ∈ *ε*}≠∅ and {*m*_0_ ∈ *ℐ*|(*m*_0_, *m*_1_) ∈ *ε*} = ∅, i.e., infector is confirmed and infectee is confirmed. Such an individual is represented as neither a root nor a leaf in a directed tree graph.

Given an infection network, one can find the following properties due to the characteristics of infectious disease transmission:

- The number of connected components with more than two vertices (individuals) equals the number of individuals (vertices) of the u-t type.
- The number of individuals excluding the u-u type represents the total sum of the number of individuals across all connected components with more than two vertices.
- The quotient of the number of individuals excluding the u-u type and the number of u-t type individuals represents the average number of individuals per connected component.
- The quotient of the number of t-t type individuals and the number of u-t type individuals represents the average number of t-t type individuals per connected component.

### 2.4 Experimental settings

Data preprocessing was performed before conducting the simulation. Firstly, 2,546 infection events (*m*_−1_, *m*_0_)∈ E were excluded due to missing report dates. Next, we identified 474 individuals, *m*_0_ ∈ *ℐ*, linked to multiple infectors, *m*_−1_∈ *ℐ*, due to uncertainty about who the actual infector is, resulting in a total of 1042 infection events, (*m*_−1_, *m*_0_) ∈ *ε* Among the identified 1042 infection events (*m*_− 1_, *m*_0_) ∈ *ε*, 480 of these cases were of the u-t type for *m*_−1_ ∈ *ℐ*. Finally, we excluded the connected components that include the u-t type from our data. Through all these preprocessing steps, the total number of confirmed cases obtained is 164,314. All simulations were done in Python version 3.9. The calculation of R_*t*_ was carried out using the Epyestim library, employing Epyestim’s default distributions and parameters. This library is described in Thompson et al. (32).

## 3 RESULTS

### 3.1 Analysis for infection network by time periods

Analyzing daily confirmed cases alone is insufficient to fully understand the transmission dynamics of infectious disease. Therefore, as depicted in Figure 2, confirmed cases have been categorized into four types, and a period analysis was conducted. In Figure 2 upper panel, the period with the highest proportion of u-u type cases among the four types was *P* 1. In contrast, the highest proportions for the remaining three types were observed in *P* 4. Moreover, the cumulative number of confirmed cases during *P* 4 shows a sharp increase, especially in the number of t-u type cases. On February 23, 2021, the cumulative number of u-t type cases surpassed that of u-u type. However, starting from April 26, 2021, the cumulative number of u-u type cases began to increase sharply. The number of cumulative confirmed cases for u-t type is almost the same as the number for t-t type over *P* 4, *P* 5.

**Figure 1.**
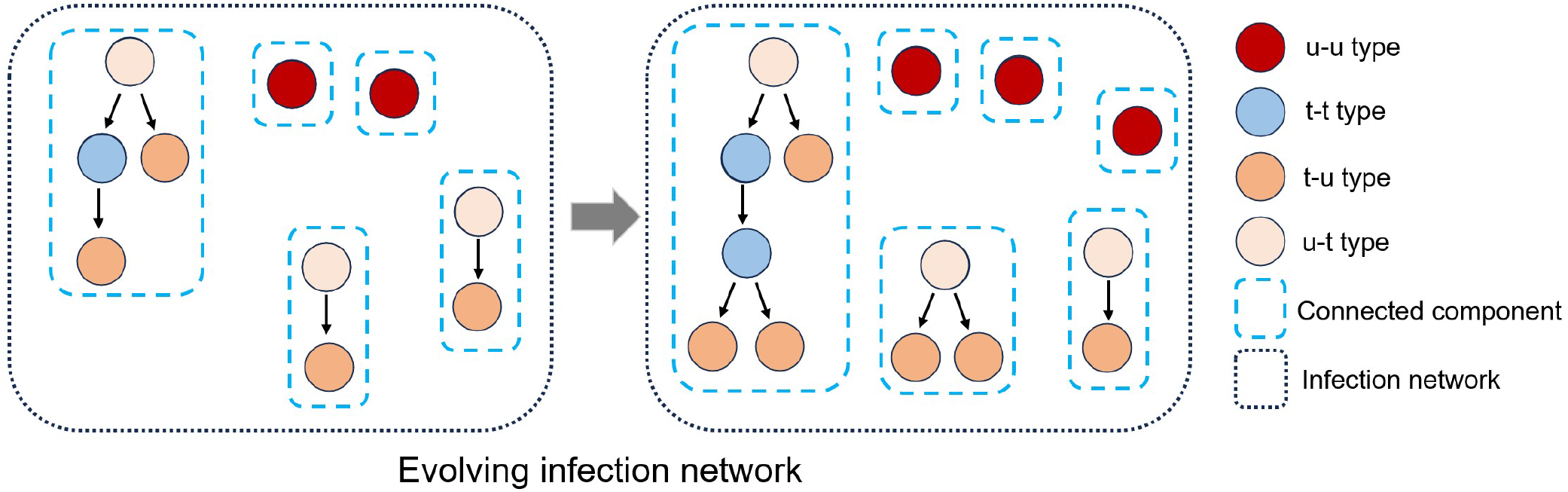
The established infection network comprises many connected components due to missing vertices (individuals) and edges(infection events). An infection network’s vertices can be classified into four types (u-t, u-u, t-u, t-t) based on the classification of their infector or infectee status as either traced or untraced. Also, the infection network evolves as an infectious disease spreads over time.

**Figure 2.**
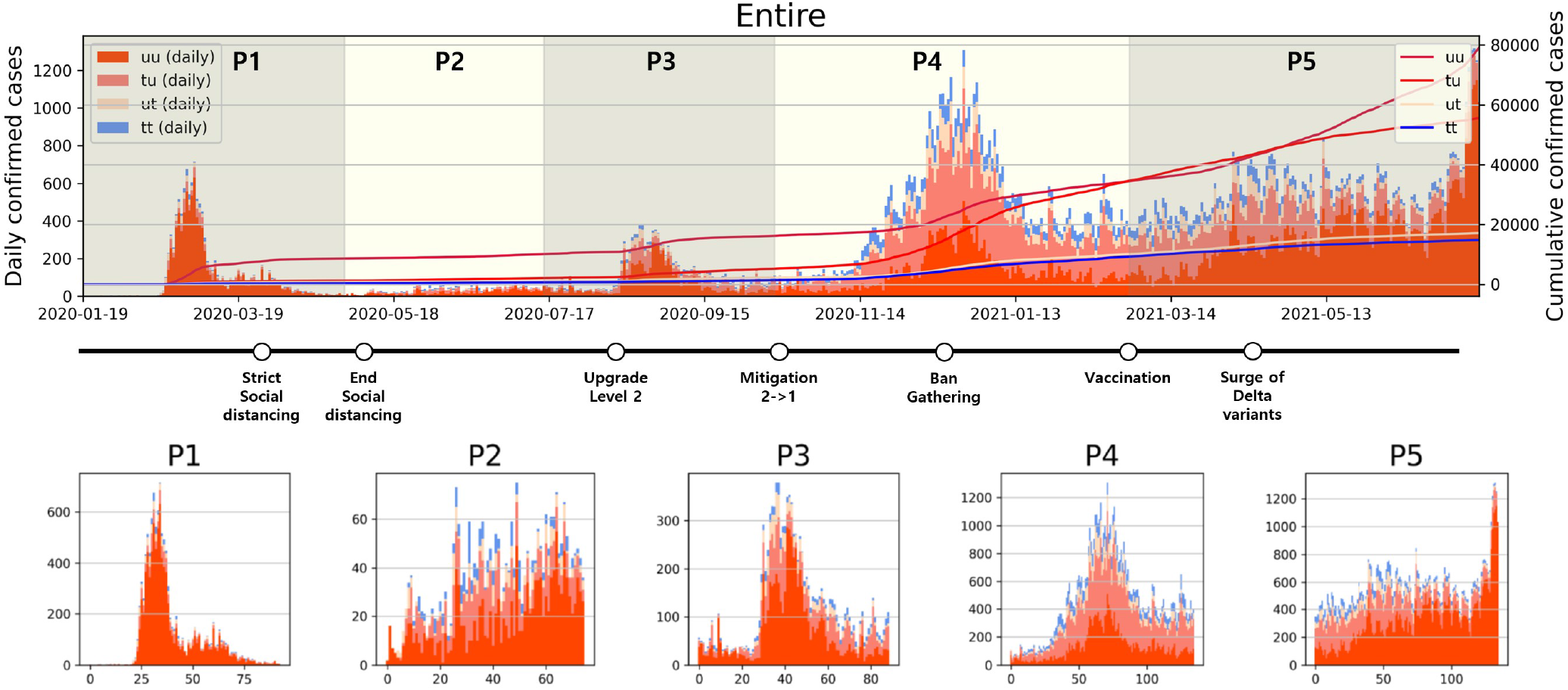
Categorized daily and cumulative confirmed cases over various periods are presented: **(Upper)** Entire period, **(Lower)** *P* 1 to *P* 5, along with representative control measures implemented in South Korea. The contrasting background colors distinguish each period.

### 3.2 Analysis for infection network by time periods and age group

The transmission dynamics might be related to the contact pattern between age groups (7, 26, 33). Figure 3 upper panel displays the age distribution of four types for both *P* 1 and *P* 4. During *P* 1, a high number of confirmed cases were observed in individuals in their 20–29 and 50–59. Among all age groups of confirmed cases, 79% were classified as the u-u type. The highest proportion of u-u type cases was found in the 20–29 age group, accounting for 88% of the cases in this age group, while the lowest was in the 0–9 age group, with 49%. However, in *P* 4, there was a distinct shift with the majority of confirmed cases being of the t-u type. This was most pronounced in the 0–9 age group, which had the highest proportion of t-u type cases at 62%, whereas the 60–69 age group had the lowest at 42%. Additionally, throughout the entire period under study, the 0–9 age group consistently exhibited the highest proportion of t-u type cases, accounting for 47%. For the age distribution in other periods, refer to Appendix Figure 7. Figure 3 lower panel presents a heatmap representing the proportion of each case type within specific age groups over the cumulative period. For instance, on the u-u type heatmap, if the y-axis is labeled 20-29 and the *x*-axis indicates 400 days (February 28, 2021), the value corresponds to the proportion of 20-29 age group cases that are classified as u-u type up to 400 days. Due to the low number of cumulative confirmed cases in the early stages of COVID-19 spread, this paper will not interpret the results for this period. When considering the entire cumulative period, the age groups with the highest proportions of u-t type and t-t type cases are 70–79 and 50–59, respectively, each accounting for 13% and 11%. The heatmaps for each type are examined in sequence. Firstly, examining the u-u type heatmap, it is observed that until the mid-period of *P* 4, the majority of confirmed cases in the 20–29 age group were of the u-u type. This trend is not exclusive to the 20–29 age group; up until the mid-period of *P* 4, a high proportion of u-u type cases is evident across most age groups. However, post the mid-period of *P* 4, there is a significant reduction in the proportion of u-u type cases in all age groups except for 20–29. Next, the t-u type heatmap shows a pattern opposite to that of the u-u type. The u-t type heatmap indicates an increase in the proportion of u-t type cases among the 40–79 age group after the mid-period of *P* 4. Lastly, the t-t type heatmap reveals an increase in the proportion of t-t type cases among the 40–69 age group posts the mid-period of *P* 4. We also analyzed the relationship between each type in terms of age group and period. As shown in Table 1, the value obtained from dividing the number of confirmed cases with traced infectors (or just traced infectors) by the number of confirmed cases with untraced infectors (or just untraced infectors) was calculated for each period and age group. In all periods except for *P* 2, the age group of 9 years and under has higher values compared to other age groups, and the 20−29 age group has the lowest values. Furthermore, this paper investigated the number of traced infectors and the number of untraced infectors across different age groups over time. These values were processed using a smoothing function with a uniform kernel of 10 points, where each point is weighted equally (1/10), to enhance data visualization and analysis. As shown in Figure 4, in *P* 4, for individuals aged 20 and above, the number of untraced infectors is almost the same as the number of traced infectors. However, in the age group below 20, there were more cases with a traced infector than with an untraced one. During *P* 5, there was a significant increase in the number of untraced infectors in the 0–59 age group.

**Table 1.** It represents the ratio of the number of traced infectors to the number of untraced infectors for each period and age group. The red (resp. blue) color stands for the age group with the maximum (resp. minimum) ratio for each period.

|  | 0–9 | 10–19 | 20–29 | 30–39 | 40–49 | 50–59 | 60–69 | 70–79 | 80–89 | 90–99 |
| --- | --- | --- | --- | --- | --- | --- | --- | --- | --- | --- |
| P1 | **0.92** | 0.21 | **0.07** | 0.20 | 0.21 | 0.18 | 0.16 | 0.17 | 0.19 | 0.16 |
| P2 | 0.95 | 0.64 | 0.45 | **0.28** | 0.46 | 0.93 | 1.16 | **1.63** | 1.13 | 1.40 |
| P3 | **0.86** | 0.77 | **0.41** | 0.44 | 0.50 | 0.50 | 0.56 | 0.57 | 0.73 | 0.48 |
| P4 | **2.68** | 2.20 | **1.16** | 1.19 | 1.38 | 1.45 | 1.31 | 1.26 | 1.79 | 2.03 |
| P5 | **0.87** | 0.61 | **0.38** | 0.44 | 0.46 | 0.53 | 0.55 | 0.59 | 0.85 | 0.80 |
| Entire | **1.32** | 0.94 | **0.51** | 0.62 | 0.67 | 0.76 | 0.77 | 0.79 | 1.14 | 1.28 |

**Figure 3.**
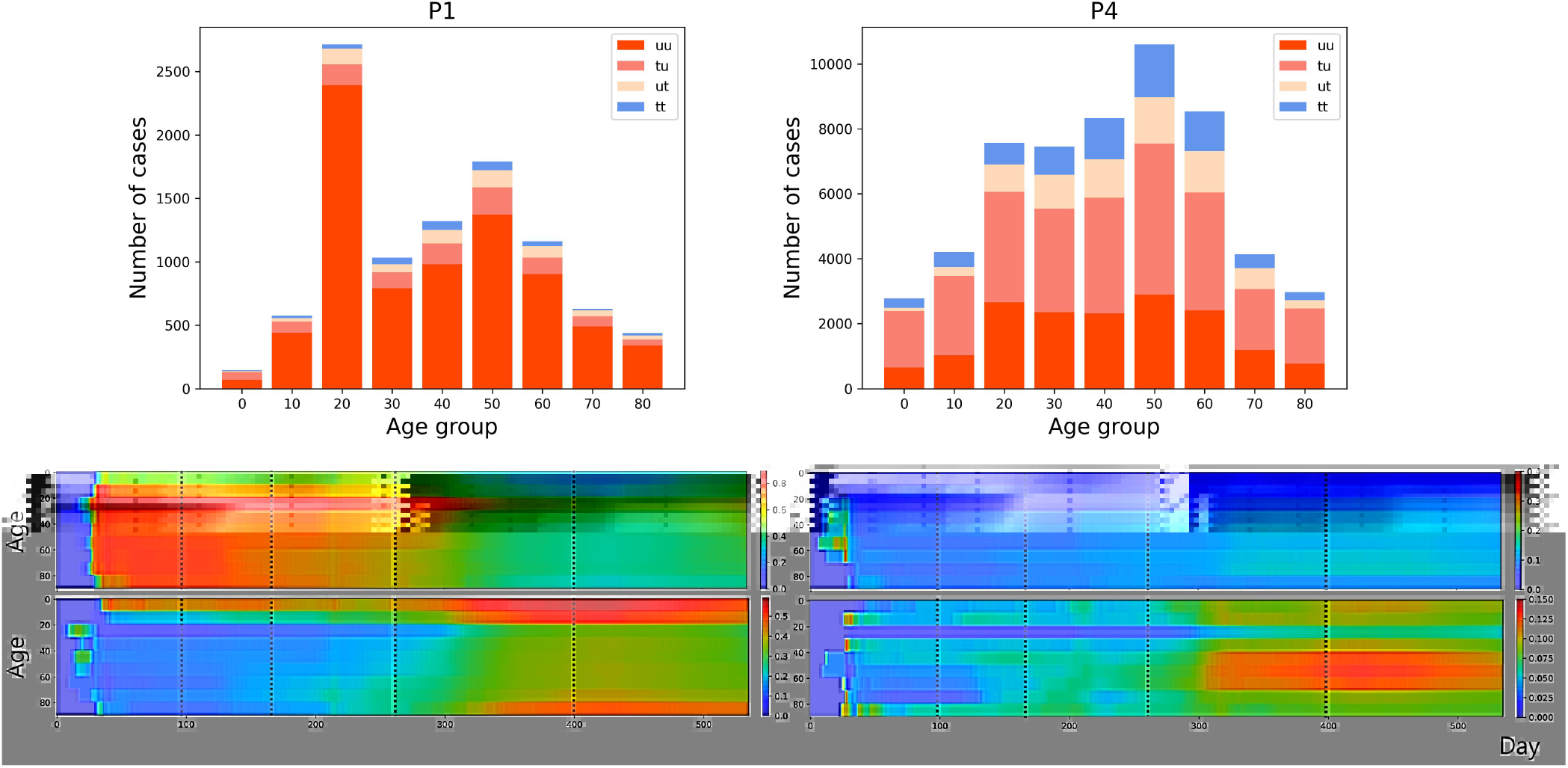
**(Upper)** Age distribution categorized according to four types for both *P* 1 and *P* 4. **(Lower)** The proportion of each case type within specific age groups over the cumulative period. The left panels display heatmap for u-u and t-u types, while the right panels show those for u-t and t-t types, with dotted lines in the figure marking the divisions between periods *P* 1 to *P* 5.

**Figure 4.**
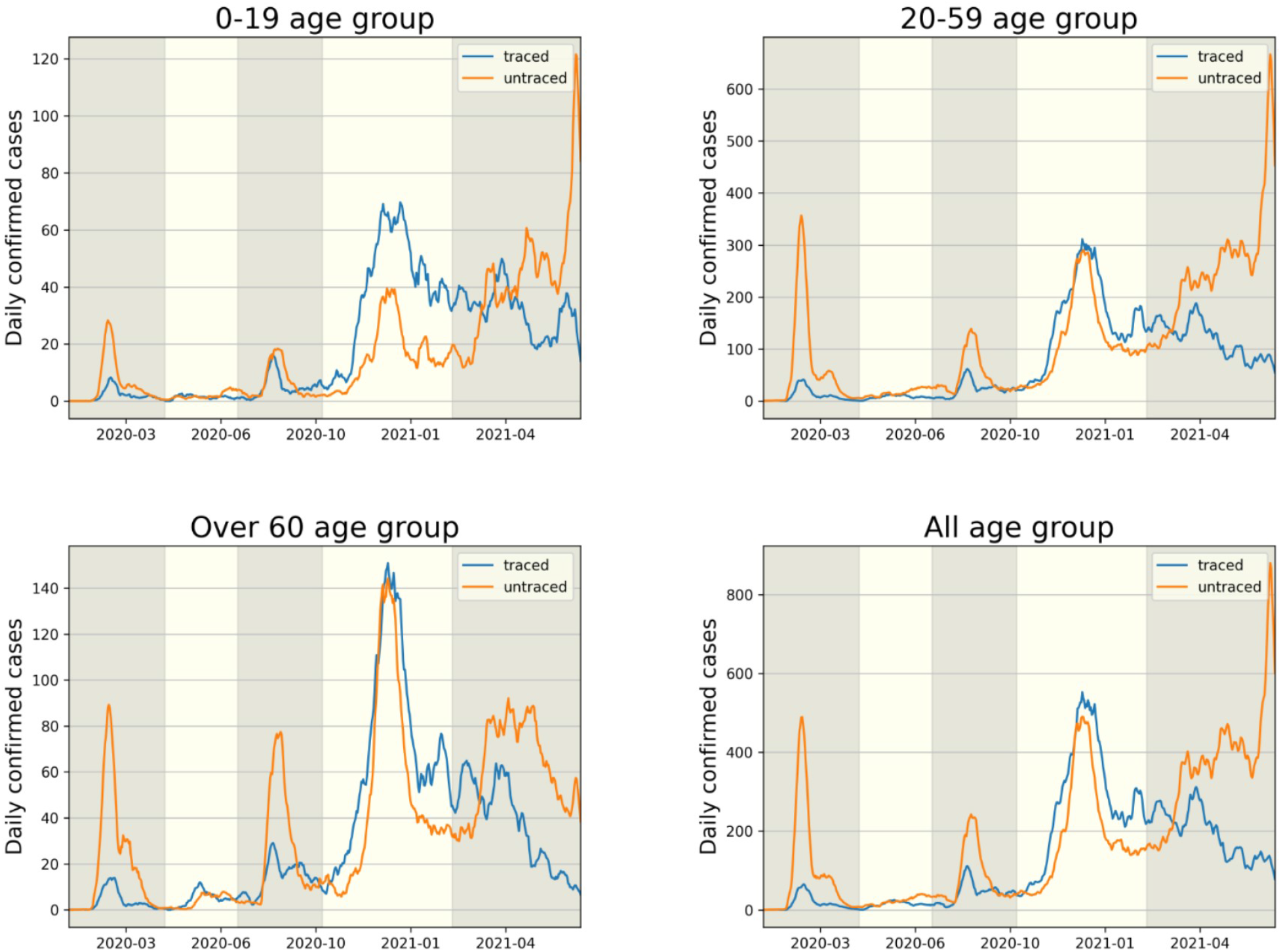
The comparison of infector identification for traced (t-u, t-t type) and untraced (u-u, u-t type) cases is shown in each age group.

### 3.3 Length of the connected components of infection network

Infection order refers to the number of subsequent infections traced back to a single confirmed case. For instance, if person A infects person B, and person B then infects person C, B and C are considered the 2nd and 3rd order infected individuals, respectively, originating from A. In this paper, we define the length of a connected component as n-1, where n is the highest order of an infector originating from a u-t type individual in the connected component. As shown in Figure 5 (Middle), in *P* 1, the proportion of connected components with a length of 1 is the highest at 81%, compared to other periods. Conversely, the lowest period is *P* 2 with 61%. For the distribution of connected component length in other periods, refer to Appendix Figure 8. In Figure 5 (Right), for the entire period and *P* 4, the slopes of the log scale for the number of cases according to length, from length=1 to lenght=2, …, and from length=8 to length=9, all exhibit similar values. Another observation is that the slope from length=2 to length=3 being closest to 0 occurs during period *P* 2. The lower panel displays the number of connected components with the length being either 1 or greater than 2, spanning the period from January 19, 2020, to July 11, 2021. During each epidemic wave *P* 1, *P* 3 and *P* 5 at their respective peaks, the number of connected components with a length of 2 or more is significantly smaller compared to the number of connected components with a length of 1.

**Figure 5.**
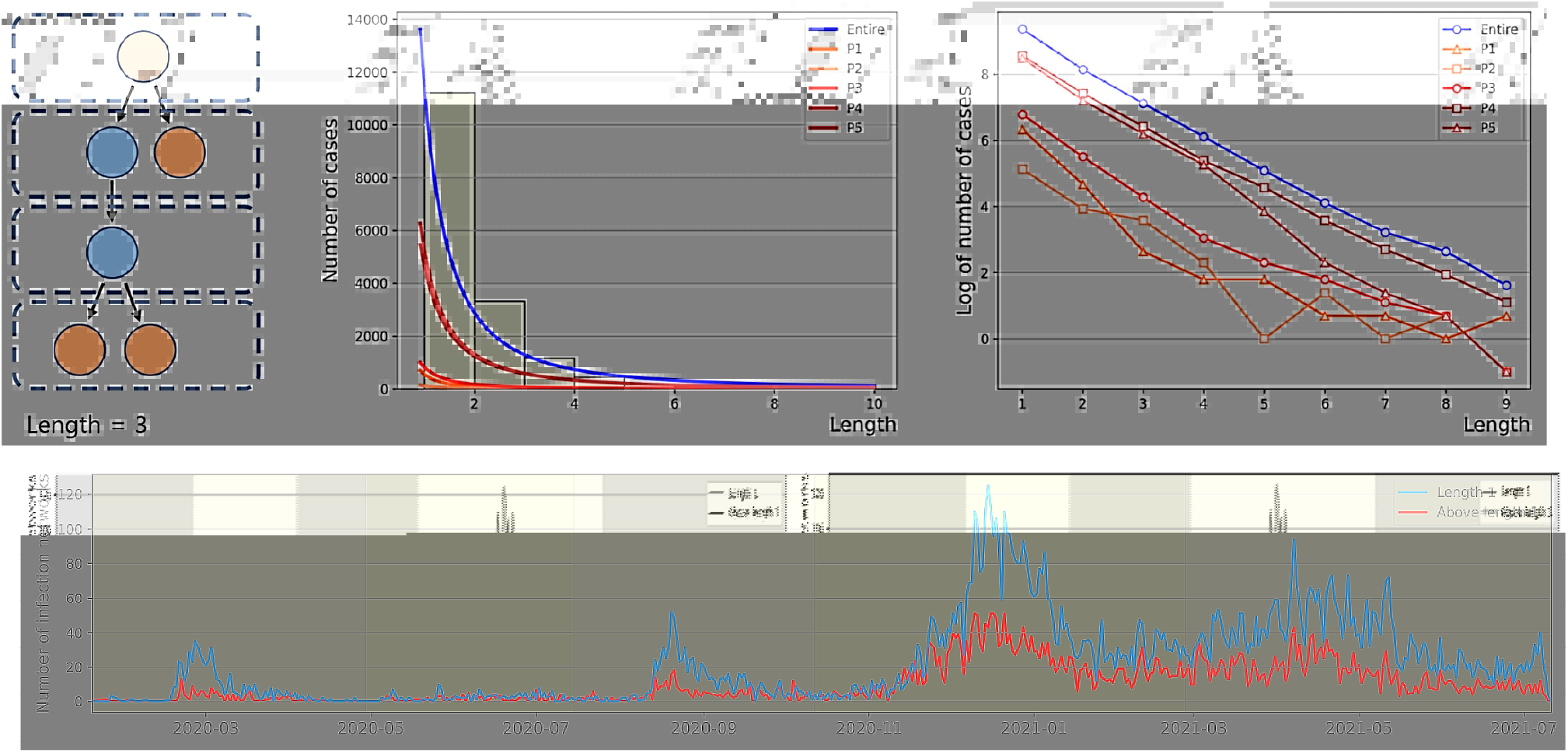
The figure upper panel presents the power law approximation of the distribution of connected component length for each period **(Middle)** and the same distributions on a log scale **(Right)**, respectively. For convenience, *y*-axis value (log value of the number of cases) of −1 indicates log 0. The lower panel represents the number of connected components by length over time.

**Figure 6.**
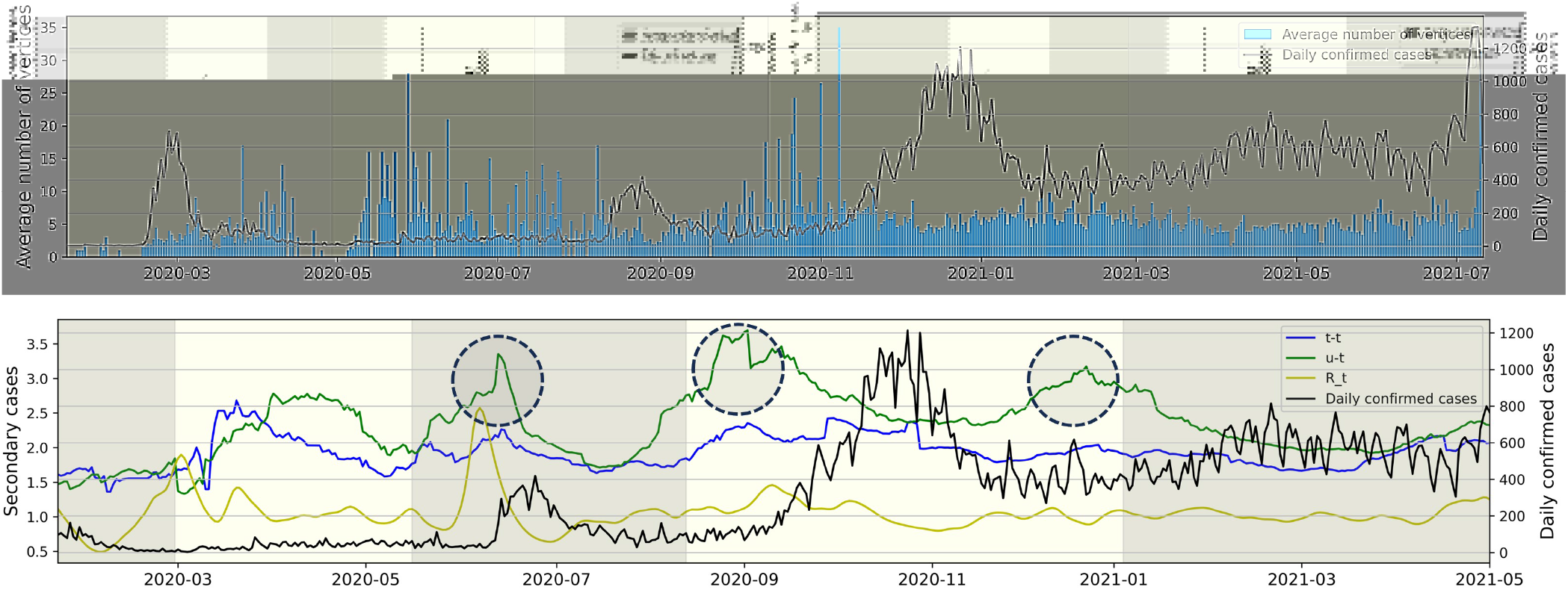
The right *y*-axis and the black line represent daily confirmed cases, while the left *y*-axis represents all other values. **(Upper)** The average number of individuals (vertices) per connected component for each day. **(Lower)** The average number of secondary cases for each type and time-dependent reproduction number ℛ_*t*_ over time.

### 3.4 Daily confirmed cases relationship

Figure 6 (Upper) represents the average number of individuals per connected component for each day from January 19, 2020, to July 11, 2021. For instance, the value for November 31, 2020, is calculated as the sum of t-t and t-u type individuals on November 31, 2020, divided by the number of u-t type individuals on the same date. The observation revealed that the value and the daily confirmed cases exhibited opposing trends. During the epidemic waves of *P* 1 and *P* 3, the value is lower compared to periods not experiencing an epidemic wave. Following the surge in daily confirmed cases in *P* 4, the value remains consistent without significant increases. Figure 6 (Lower) illustrates the average number of secondary cases for both u-t and t-t types, calculated with a window size of 30, from March 22, 2020, to July 11, 2021, and also depicts the time-dependent reproduction number R_*t*_ (12). The value is an indicator derived from the infection network analysis. For instance, the average number of secondary cases for the u-t (resp. t-t) type on August 1, 2020, is defined as the real-time calculated average value of confirmed cases directly infected by the u-t (resp. t-t) type within the infection network identified between July 1, 2020, and August 1, 2020. For instance, if within the identified infection network for the period, there are 3 connected components, and the number of individuals infected by each u-t type individual is 2, 6, and 1, respectively, then the average number of secondary infections for the u-t type on August 1, 2020, is calculated as (2+6+1)/3=3. The time-dependent reproduction number R_*t*_ did not show a significant increase before an increase in daily confirmed cases during *P* 4 and *P* 5. However, the circular markers in Figure 6 (Lower) indicate a significant increase in the average number of secondary cases for u-t type.

## 4 DISCUSSION

Despite having a large volume of epidemiological data due to its active contact tracing efforts compared to other countries, South Korea’s infection network, generated from the data, comprises many connected components as a result of numerous missing vertices (individuals) and edges (infection events). This article analyzed the infection network using vertices of four types: u-u, u-t, t-u, and t-t based on whether their infector or infectee falls into the traced or untraced category. We analyzed the dynamics of the infection network based on each type, time, and age group, deriving insights. Our results showed a significant surge in the number of t-u type cases (i.e., traced infector - untraced infectee type) during *P* 4 when the government upgraded the social distancing level twice as well as expanding the screening clinics in Figure 2. A significant surge in the cumulative number of u-u type cases was also observed, beginning in the mid-phase of *P* 5, coinciding with the spread of the Delta variant. The average number of t-t type individuals per connected component close to 1 in *P* 4 and *P* 5 indicates active contact tracing in response to mass infection. In other words, the proposed method allows for the analysis and evaluation of phenomena induced by various events such as the implementation of public health policies, the emergence of new variants, and more.

Our result also found age-specific transmission patterns for the four types in Figure 3. Individuals of the u-u type pose a significant risk of causing mass infections in the community. Across periods *P* 1 to *P* 5, the highest proportion of u-u type cases (57.4%) was observed in the 20–29 age group. This can be inferred to be due to the 20–29 age group’s wider range of activities and frequent interactions with various people. The 0–9 (47.6%), 10–19 (40.9%), and 80–89 (46.5%) age groups had the highest rates of t-u type cases, indicating these demographics may serve as key points for interrupting transmission chains. By focusing on these patterns in the implementation of public health policies, it may be possible to more effectively contain outbreaks and prevent wider community spread. Individuals of the u-t type, as initial infectors in a connected component, help identify which age groups had more asymptomatic COVID-19 cases and were more engaged in contact tracing, based on their age-wise proportions. Across periods *P* 1 to *P* 5, the highest proportion of u-t type cases (13%) was observed in the 70–79 age group. From mid *P* 4, it was observed that the proportion of u-t type cases in the 30-79 age group was higher compared to other age groups. The proportion of t-t type cases by age group also allows for the inference of which age groups were more actively involved in contact tracing. Across periods *P* 1 to *P* 5, the highest proportion of t-t type cases (11%) was observed in the 50–59 age group. After mid *P* 4, the 40–69 age group showed a higher proportion of t-t type cases compared to other age groups. Furthermore, the analysis of the value obtained from dividing the number of confirmed cases with traced infectors (or just traced infectors) by the number of confirmed cases with untraced infectors (or just untraced infectors) across age groups revealed a sequence of 0–9 *>* 90–99 *>* 80–89 *>* 10–19 *>* 70–79 *>* 60–69 *>* 50–59 *>* 40–49 *>* 30–39 *>* 20–29. For the 0–9 and 80–99 age groups, where the number of contacts is limited, contact tracing was more manageable; however, in age groups like 20–39, which have a higher number of contacts, contact tracing was found to be more challenging. These analyses provide valuable information for understanding the transmission dynamics of COVID-19, allowing us to suggest strengthening or relaxing control measures for specific age groups based on the period’s characteristics.

Our results also investigated the distribution of the lengths of connected components within the infection network. In *P* 2, the proportion of connected components with a length of 1 was the lowest, while the proportions with lengths of 2 and 3 were the highest. This indicates that during *P* 2, which had the lowest daily average of 37 confirmed cases, the infection network had fewer missing edges (infection events). Further investigation across the entire period, as shown in the lower panel of Figure 5, revealed an increase in the number of connected components with a length of 1 during surges in daily confirmed cases. The earlier results motivated the hypothesis that the average number of individuals per connected component for each day would decrease during spikes in infections. This was indeed observed in the upper panel of Figure 6. It means that when the number of daily confirmed cases surges, it becomes challenging to contact trace high-order transmissions. This phenomenon may stem from changes in the government and the public’s willingness to engage in contact tracing and limitations of existing contact tracing methods in the face of a highly infectious virus spreading worldwide. For this reason, this article proposed the average value of confirmed cases directly infected by the u-t type as an indicator of infectious disease transmission potential. Utilizing the infection network up to 30 days prior allows for real-time calculation, and this indicator shows high values before a surge in daily confirmed cases. Due to the indicator allowing for an approximation of real-time unreported cases, it is more sensitive compared to R_*t*_ and increases before the third epidemic wave. We anticipate it to be a useful indicator in situations like in South Korea, where active contact tracing is conducted.

Our study has several limitations. Firstly, we do not consider unreported cases including asymptomatic individuals, those with mild symptoms who were not tested, and unreported self-tests from the surveillance pyramid (34). Considering unreported cases is a key research topic for understanding and predicting the scale of infections (35, 36, 37). Acknowledging the constraints imposed by unreported cases, especially concerning COVID-19 transmission within contact networks, we recognize the potential of methods such as multiple imputation techniques (35) and data augmentation through link prediction (36) to provide valuable insights. Furthermore, the exploration of machine learning-based approaches (37) presents another promising avenue for addressing data gaps. Studies that have not estimated unreported cases but have specifically limited unreported cases to environmental factors include Myall et al. (38), which analyzed patient-contact networks using patient contacts obtained from hospital health records. Despite its limitations, the KDCA data we analyzed remains trustworthy. According to the KDCA, based on serological surveillance and contact tracing data, the rate of unreported cases in South Korea from January 19, 2020, to July 30, 2022, was approximately 19.5%. This rate is notably lower than those seen in international contexts, a difference attributed to the widespread availability of testing and the public’s adherence to control measures (39, 40). Secondly, our study did not quantitatively assess contact tracing effectiveness. There are several previous studies about the effectiveness of contact tracing strategies for COVID-19 (41, 42, 1). Kretzschma et al. (41) analyzed contact tracing effectiveness using a stochastic model, finding that immediate tracing and testing are crucial for reducing the spread of COVID-19. Delays in testing and tracing significantly diminish the potential to keep the effective reproduction number below 1. Korean government implemented the contact tracing described in (42). Contact tracing for COVID-19 was performed using information from credit card records, handwritten visitor logs, QR codes through KI-Pass, and the Safe Call system after interviews in Korea. Hellewell et al. (1) found tracing and isolation could control outbreaks within 12 weeks. There are previous studies to investigate the infection network of COVID-19 in (2, 43, 44). Luo et al. (43) in 2021 developed an infection network considering the history of exposure and transmission source. The visualization method, which identifies vertices in the infection network as clusters of infected individuals, revealed a highly central infection cluster in (44). However, we developed an infection network, categorizing infector-infectee pairs by age group and periods, specifically focusing on untraced cases. Jo et al. (2) emphasized the importance of gathering network data and examining network structures to improve the effectiveness of governmental responses to COVID-Additionally, in future research, we intend to expand our analysis to encompass infection networks incorporating spatial information, as discussed in (45).

The current research reveals that, despite active contact tracing efforts, South Korea’s infection network, derived from a large volume of epidemiological data, comprises many connected components due to numerous missing entities (individuals) and infection events (edges). The presence of numerous connected components complicates the inference of relationships between vertices. Therefore, a four-type classification method for vertices (confirmed cases) is proposed. This method enables the categorization of vertices within the numerous distinct connected components from a common perspective, thereby facilitating the analysis and interpretation for each vertex type. The changes in the number of cases for each type over time relate to the emergence of new coronavirus variants (such as Delta) or the implementation of control measures. When analyzed by age group, it was observed that certain age groups are more sensitive to these events. Additionally, we analyzed the infection network from the perspective of connected components, proposing a new indicator and comparing it with R_*t*_. Despite limitations, the study’s categorization of epidemiological data into four types not only offers a robust foundation for evaluating public health policies and comprehending the dynamics of COVID-19 transmission but also serves as a foundational health planning tool for resource management and tool selection/development for contact tracing.

In conclusion, South Korea’s epidemiological data generated from active contact tracing enables novel infection network analysis. Our analysis reveals significant age-specific transmission patterns, particularly in the 20–29, 40–69, and 0–9 age groups. The patterns show a distinct shift around the midpoint of *P* 4, with the 20–29 (57.4%) age group exhibiting the highest proportion of u-u type cases, the 40–69 age group predominantly showing u-t and t-t types, and the 0–9 (47.6%) age group having the highest rate of t-u type cases across entire periods. This suggests a relationship between age groups and the four-type classification. A significant increase in t-u and u-u type cases was observed during certain periods, providing opportunities for analysis and evaluation of phenomena induced by various events, such as the implementation of public health policies, the emergence of new COVID-19 variants, and more. Also, through the investigation of the distribution of lengths of connected components within the infection network, we found that the average number of individuals per connected component tends to decrease during surges in daily confirmed cases, indicating that tracing high-order transmissions becomes more challenging. Accordingly, we propose the average value of confirmed cases directly infected by the u-t type as an indicator to assess the potential for infectious disease transmission. Additionally, this approach could facilitate the early detection of changes in willingness among individuals to participate in tracing, or in the reduced capacities of contact tracing systems. The investigation of infection networks is crucial for advancing our capacity to control and mitigate the transmission of infectious diseases. Recognizing the necessity for a more thorough age-based categorization, the study emphasizes potential areas for future research improvements in comprehending and refining public health strategies. Additionally, our study presents a new real-time indicator using contact tracing data collected during actual infection spread, ultimately providing support for decision-makers and contributing to reducing the pandemic’s impact on global communities.

## Data Availability

Epidemiological data were obtained from Korea Disease Control and Prevention Agency (KDCA). The data is openly available.

https://www.kdca.go.kr/

## DATA AVAILABILITY STATEMENT

The original contributions presented in the study are included in the article material, further inquiries can be directed to the corresponding author.

## AUTHOR CONTRIBUTIONS

HW, Lee and HY, Choi: analyzed the data. HW, Lee,HY, Choi, HJ, Lee, SM, Lee and CH, Kim: drafted and revised the manuscript. HJ, Lee, SM, Lee and CH, Kim: interpreted the results. All authors contributed to the article and approved the submitted version.

## FUNDING

This work was supported by the National Research Foundation of Korea(NRF) grant funded by the Korea government(MSIT) (No. 2022R1A5A1033624).

## ACKNOWLEDGMENTS

Epidemiological data were obtained from Korea Disease Control and Prevention Agency (KDCA) (27).

## CONFLICT OF INTEREST

The authors declare that the research was conducted in the absence of any commercial or financial relationships that could be construed as a potential conflict of interest.

## APPENDIX

**Figure 7.**
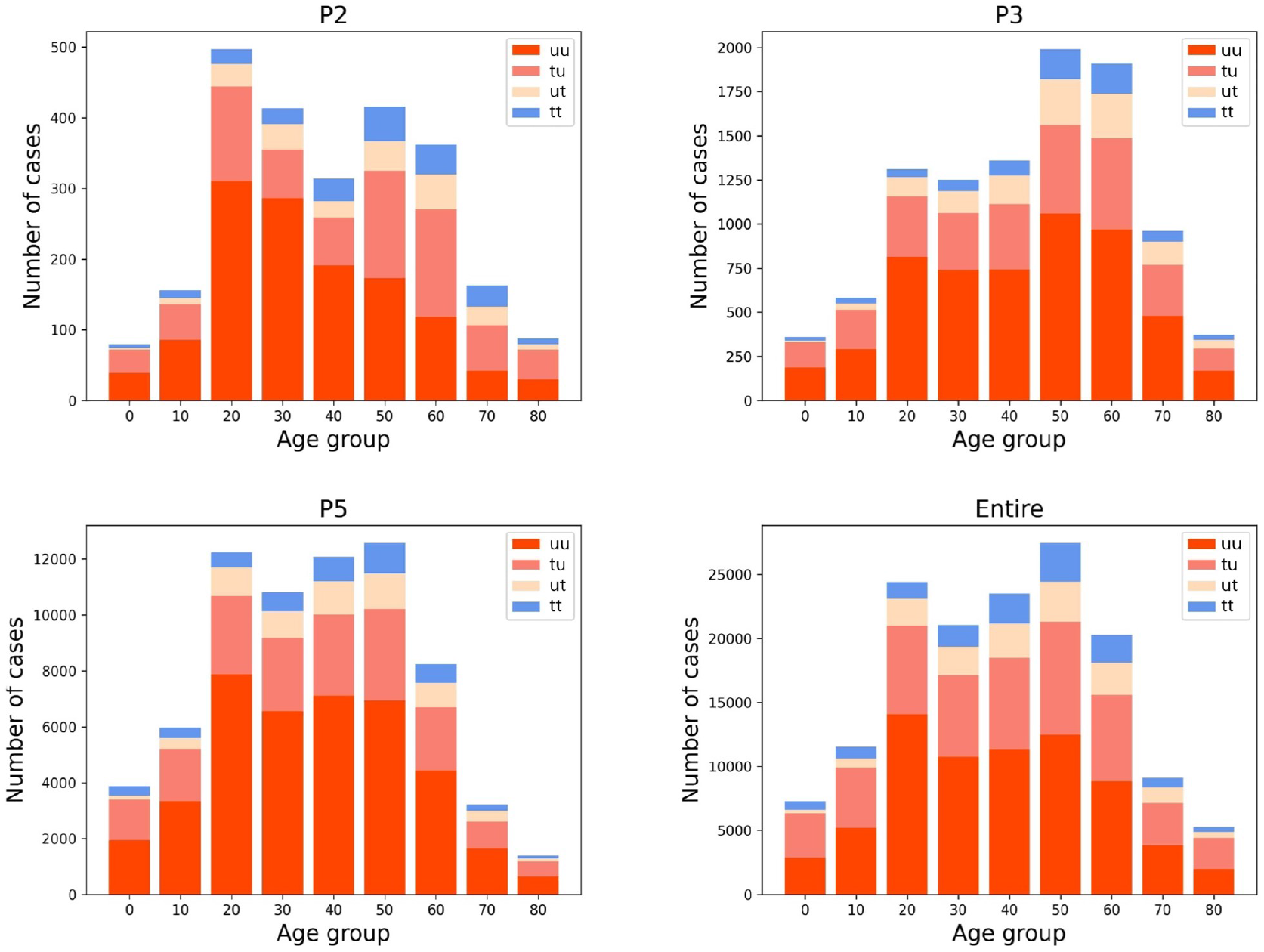
Additional information for Figure 3. Age distribution categorized according to four types for both *P* 2, *P* 3, *P* 5 and Entire.

**Figure 8.**
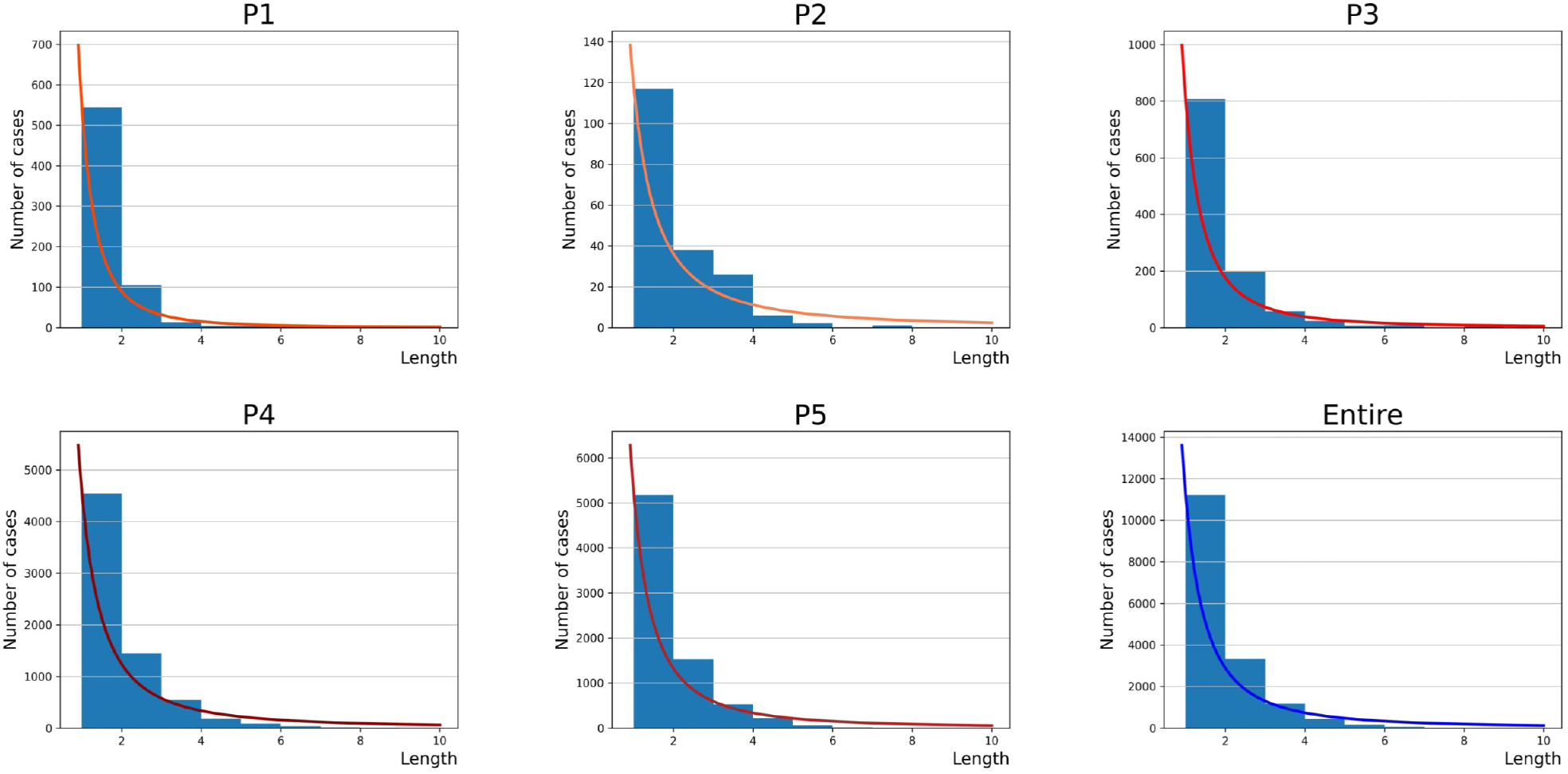
Additional information for Figure 5. The figure presents the distribution of connected component length for each period.

## Notes

### Competing Interest Statement

The authors have declared no competing interest.

